# The Brazilian vaccine divide: how some municipalities are being left behind in the Covid-19 vaccine coverage

**DOI:** 10.1101/2023.05.23.23290401

**Authors:** Antonio Fernando Boing, Alexandra Crispim Boing, Lorena Barberia, Marcelo Eduardo Borges, SV Subramanian

**Author notes:** **Correspondence** Antonio Fernando Boing, Federal University of Santa Catarina, Florianópolis, Brazil, 88040-900.

## Abstract

**Objectives:** This study aims to assess the progress of geographic, socioeconomic, and demographic disparities in Covid-19 vaccination coverage in Brazil over the first two years of the vaccination campaign.

**Study design:** Ecologic study.

**Methods:** Data from the National Immunization Program Information System were used to estimate covid-19 vaccine coverage. Brazilian municipalities were divided into two groups based on their vaccine coverage for the booster dose. The first group comprised 20% of municipalities with the lowest coverage, while the second group (80% of municipalities) had higher coverage. The analysis was conducted separately for four age groups: 5-11, 12-17, 18-59, and 60+. Exploratory variables included socioeconomic and health services indicators. Crude and adjusted logistic regression models were used to estimate the probability of a municipality being among those with the worst vaccination coverage according to the categories of exploratory variables.

**Results:** Between January/2021 and December/2022, Brazil administered 448.2 million doses of the covid-19 vaccine. The booster vaccination coverage varied from 24.8% among adolescents to 79.7% among the elderly. The difference between the group with the highest and lowest coverage increased during the national vaccination campaign. Municipalities with lower education levels, higher proportion of Black population, higher Gini index, and worse health service indicators had a greater likelihood of having lower vaccination coverage.

**Conclusions:** High and increasing levels of inequality in Covid-19 vaccination were observed in Brazil across all age groups during the vaccination campaign in 2021-2022.

## Introduction

By April 2023, over 764 million cases and nearly 7 million deaths from COVID-19 have been reported globally and Brazil remains one of the countries most severely impacted by the pandemic.^1^ The fight against covid-19 in Brazil from 2020 to 2022 was characterized by a lack of central coordination and the failure to utilize the best scientific evidence to guide public policies.^2-3^

Despite ample evidence on the effectiveness, safety, and cost-effectiveness of Covid-19 vaccines,^4-5^ the vaccination rollout in Brazil has been notably sluggish. While initial dose coverage for the elderly and adults exceeds 85%, there is a significant shortfall in booster doses and overall coverage for adolescents, children, and infants.^6^ Furthermore, disparities in vaccination coverage have emerged at subnational levels, particularly among adults and the elderly,^7^ reflecting the pre-pandemic inequalities observed in influenza,^8^ measles,^9^ and polio^10^ vaccination efforts in Brazil.

Numerous countries globally have expressed concerns regarding disparities in vaccine coverage, highlighting significant social, economic, and racial/ethnic inequalities.^11-12^ In the case of Brazil, the situation is particularly alarming, despite the presence of a public, universal, and comprehensive healthcare system, due to the country’s profound social and income inequalities, ranking among the highest worldwide.^13^ Challenges are amplified by factors such as inadequate sanitation and housing conditions^14^, disparities in healthcare access^15^, and a high prevalence of chronic diseases.^16^ Moreover, the government’s inadequate and fragmented pandemic response, the rampant dissemination of anti-vaccine misinformation, disincentives to vaccination, and increasing vaccine hesitancy have further exacerbated the situation.^17^

The global and national impact of the pandemic underscores the need for continuous evaluation of inequalities in COVID-19 vaccination efforts. This requires conducting comprehensive national studies, covering extended periods, and employing finer geographic disaggregation. Thus far, research on the Brazilian experience has predominantly focused on the early stages of the vaccination campaign^18^, specific population groups^19^, or aggregated data at the state level.^20^ Additionally, studies in Brazil and worldwide have primarily examined vaccination efforts within short time intervals, such as a single month or epidemiological week, with a need for more literature analyzing the dynamic evolution of inequalities - and countries’ responses -throughout the entire vaccination campaign.

This study aims to examine the progression of geographic, socioeconomic, and demographic disparities in COVID-19 vaccination coverage across municipalities in Brazil during the initial two years of the vaccination campaign.

## Methods

### Vaccination data

To estimate vaccination coverage against Covid-19 in Brazil, we utilized data from the National Immunization Program Information System (SI-PNI), which is available through the openDataSUS platform (https://opendatasus.saude.gov.br/). openDataSUS is an open data platform created by the Ministry of Health in Brazil that provides microdata from various health information systems.

Records were removed from the database if (1) there was no anonymized identifier for the individual, (2) there were more than six vaccine records per identifier, (3) dates of reported administration of doses were inconsistent, or if (4) information on sex, place, and date were missing or incomplete. The population vaccinated was divided into the following age groups: 5 to 11, 12 to 17, 18 to 59, and 60 years or older. The total doses administered between January 2021 and December 2022 were grouped according to epidemiological week, gender, age group, type of dose, and municipality of residence. Information on vaccination coverage is available at https://github.com/covid19br/dados-vacinas.

### Outcome

We created a dichotomous variable by categorizing the 5,570 Brazilian municipalities into two groups based on their vaccine coverage of the third booster dose during the week when Brazil reached 20% coverage in the corresponding age group. The first group included the 20% (n=1,114) of municipalities with the lowest vaccine coverage, while the second group comprised the remaining 80% with the highest coverage. For pediatric vaccination, we considered the second dose. The analysis was conducted separately for different age groups. Brazil reached 20% vaccination coverage of the analyzed dose in week 43 of 2021 among the elderly, week 4 of 2022 among adults, week 33 of 2022 among adolescents, and week 13 of 2022 among children.

### Exploratory variables

We included three exploratory variables that captured socioeconomic and demographic characteristics: income concentration (as measured by the GINI index), expected years of schooling at 18 years of age, and proportion of the black and brown population. These indicators were calculated for each of the 5,570 Brazilian municipalities using data from the 2010 Brazilian census, which is the most recent census with available data (http://www.atlasbrasil.org.br/). The Gini index is a numerical indicator based on the Lorenz curve that measures income inequality in a population. The expected years of schooling at age 18 reflects the average number of years that a child entering school will complete by age 18, based on the current patterns of school attendance. The proportion of the black and brown population was calculated based on the self-classification of the census population into five standardized categories established by the Brazilian government: Blacks and Browns (Blacks), whites, indigenous peoples, and Asians.

Municipal health services variables were: per capita expenditure on health, number of nurses plus physicians per 1,000 inhabitants, and primary health care ambulatory office per 1,000 inhabitants. The numbers of health professionals and offices were obtained from the National Register of Health Establishments (CNES). In Brazil, registration with the CNES is mandatory for all physical and professional units that provide healthcare services, such as offices, clinics, hospitals and laboratories, whether public or private. Its data are made available openly by municipality by the Department of Informatics of the Brazilian Unified Health System (https://datasus.saude.gov.br/). In the analysis, we considered the structure installed and the supply of professionals in January 2021, the month in which the covid-19 vaccination campaign began. Health expenditure data considered the municipality’s total expenditure on public health programs and services in 2020, as published by Vieira et al^21^. based on data from the Public Health Budget Information System (SIOPS), managed by the Ministry of Health (https://www.gov.br/saude/pt-br/acesso-a-informacao/siops). All exploratory variables were categorized into quartiles.

### Data analysis

For each group of the outcome variable, vaccination coverage was calculated for all epidemiological weeks up to December 2022, and the median, minimum value, 25th percentile, and differences in medians between the groups were estimated for the first and last analyzed months of each age group. The proportion of municipalities classified among the 20% of the country with the lowest vaccination coverage was calculated for each quartile of the exploratory variables and each state. Crude and adjusted logistic regression models were used to estimate the probability of a municipality being among those with the worst vaccination coverage according to the categories of exploratory variables. The adjusted model included all variables. Finally, the municipalities were plotted on maps with colors depending on whether the city has the highest or lowest vaccination rates for each respective age group. All analyses were conducted for the four age groups included in the study, and Stata 15.1 was used for the analyses, while QGIS 3.3 was used for creating the maps.

## Results

Between January 2021 and December 2022, Brazil administered 448.2 million first, second and third doses to the population aged five years or older. Vaccination coverage of the booster dose in December 2022 was 79.7%, 52.3% and 24.8% among elderly, adults and adolescents, respectively. Among children aged 5 to 11, 46.2% had taken the initial two doses. More details on vaccination coverage and socioeconomic, demographic and health service characteristics are presented in Supplementary Table 1.

Throughout the vaccination campaign, municipalities with higher initial vaccination coverage consistently maintained higher coverage than those with lower initial coverage across all age groups (Figure 1). Differences in coverage became apparent early on, with certain municipalities showing lower coverage rates than others. While both groups saw an increase in the proportion of vaccinated individuals over time, a subset of municipalities consistently reported significantly lower coverage rates than the national average. For example, when the national average coverage for the second dose reached 45.0% among children, the municipalities with the lowest coverage only administered doses to 17.2% of this age group. Table 1 demonstrates that as the vaccination campaign progressed, the difference between the medians of the groups with the highest and lowest vaccination coverage increased for children, adolescents, and adults. Among adults, when Brazil reached 20% coverage for the third dose, the median vaccination coverage among the municipalities with the lowest immunization was 8.5%, compared to 21.0% in the group of municipalities with the highest vaccinated population (a difference of 12.5 percentage points (pp)). In December 2022, these values were 33.4% and 58.7%, respectively, representing a difference of 25.3 pp. Significant disparities in vaccination coverage were consistently observed among Brazilian municipalities throughout all months, with some consistently having lower coverage than others.

**Figure 1.**
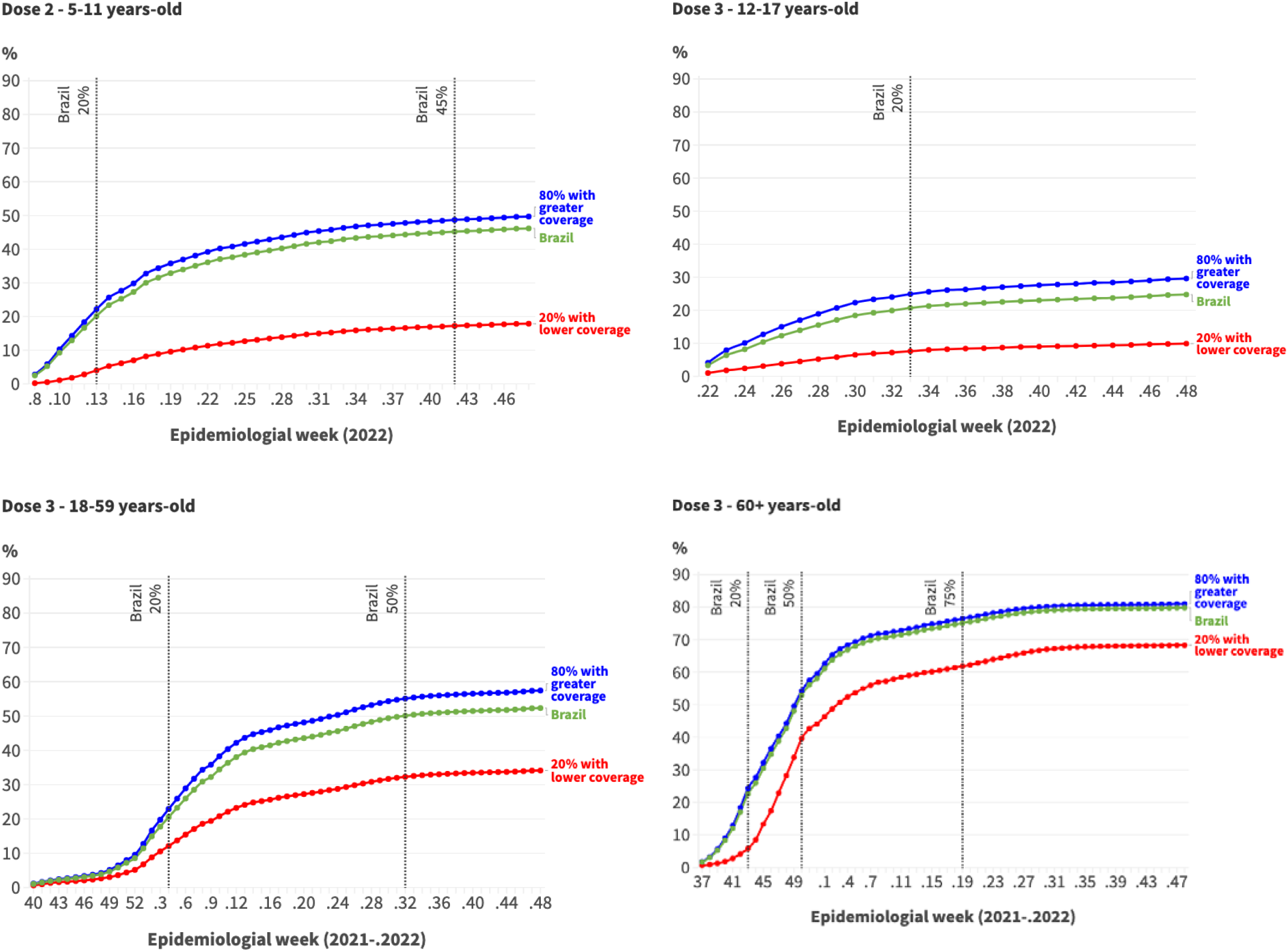
Evolution of COVID-19 vaccination coverage according to groups of municipalities with the highest and lowest proportional vaccination at the beginning of the campaign*. Brazil, 2021-2022.

**Table 1:**
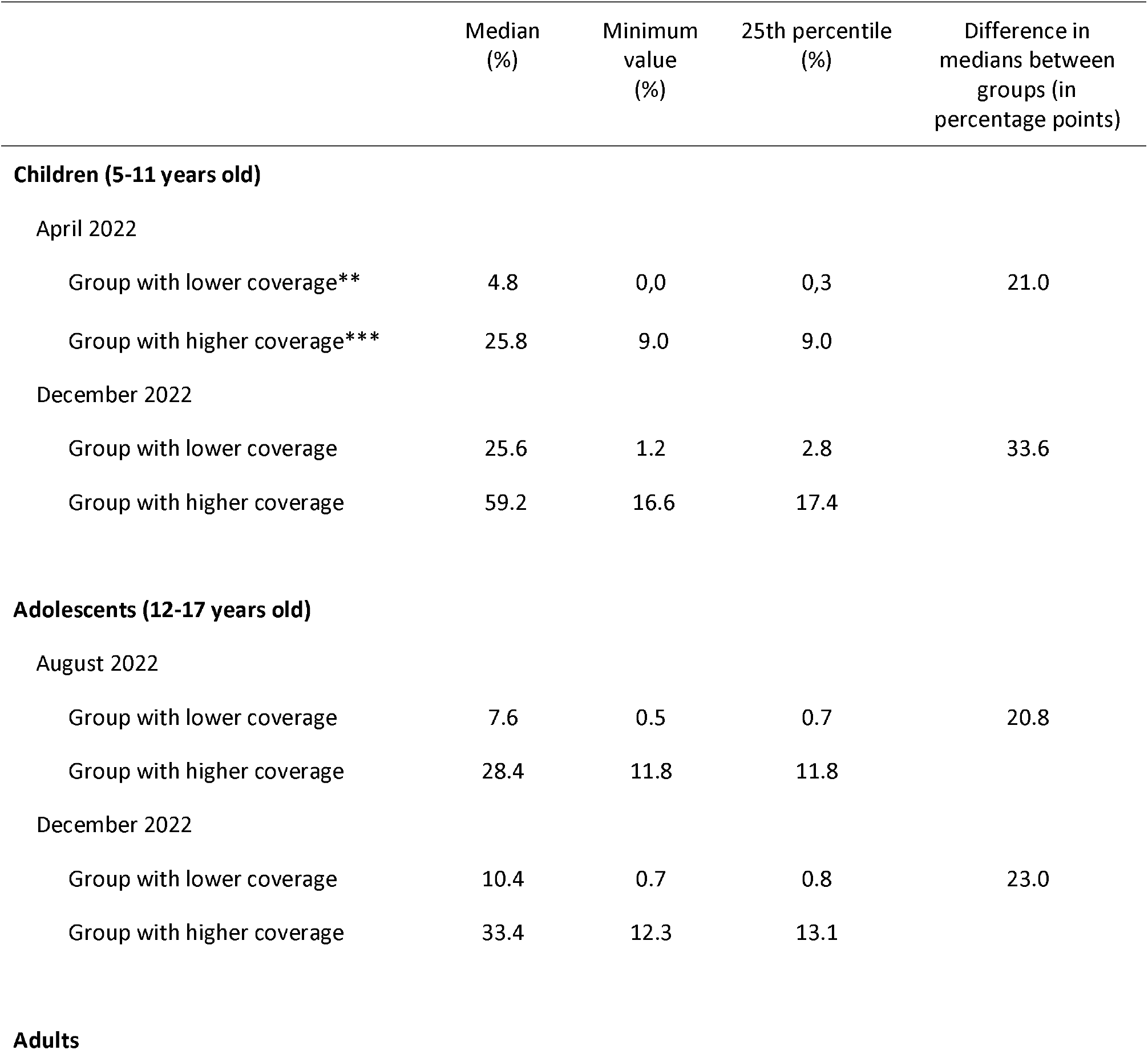
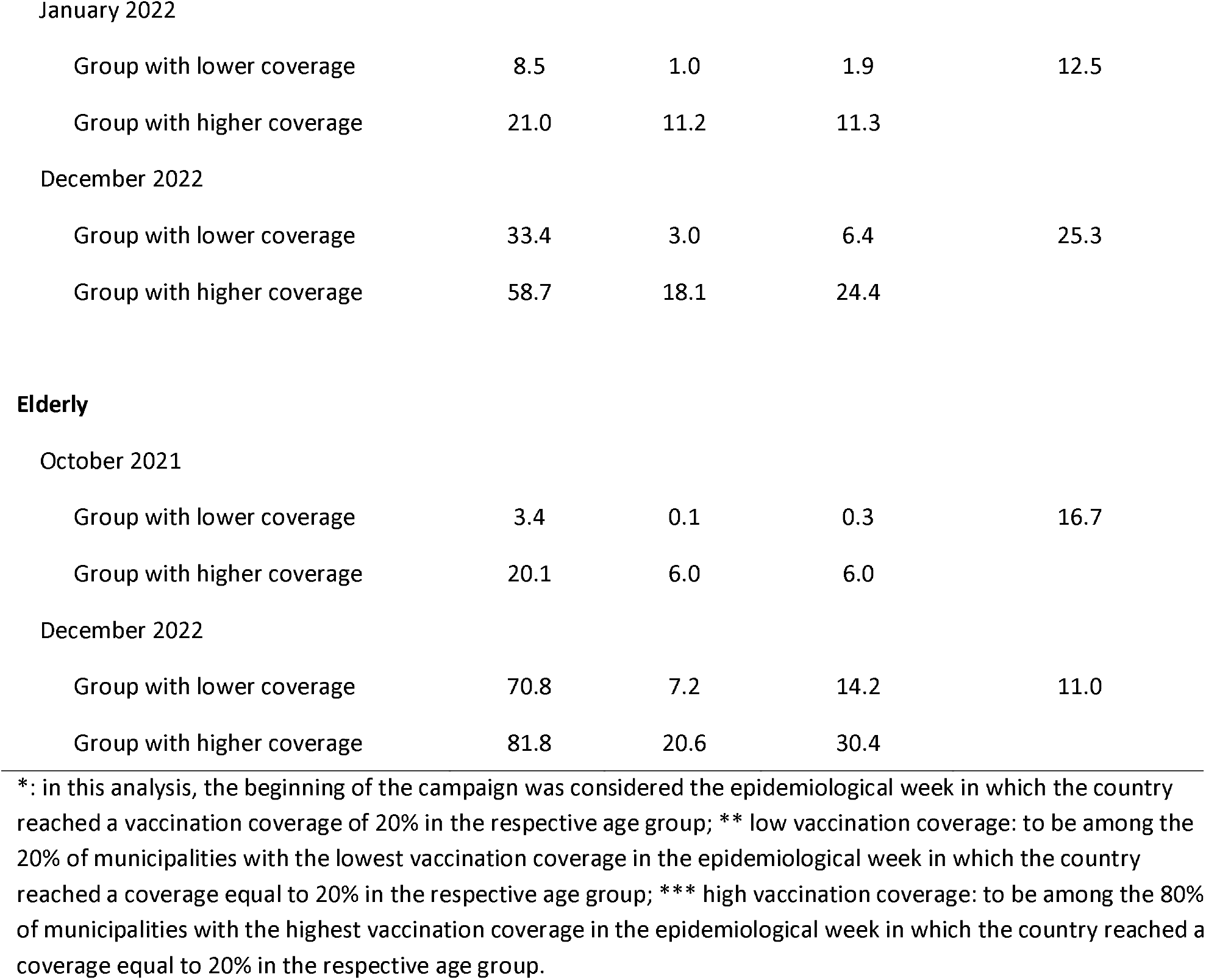
Median, minimum value, and 25th percentile of COVID-19 vaccination coverage at the beginning* of vaccination campaigns for each age group and in December 2022. Brazil, 2021-2022.

The analysis of socioeconomic, demographic, and health service characteristics of the municipalities that fell into the 20% with the lowest vaccination coverage in Brazil revealed that, in general, these municipalities have worse indicators and a higher proportion of black population (as presented in Table 2). For instance, while approximately 10% of the municipalities in the quartile with the lowest concentration of income were among those with the worst immunization coverage, the proportion was greater than one in three municipalities in the quartile with the highest concentration of income. Similar disparities were observed in other indicators analyzed.

**Table 2.** Proportion of municipalities classified among the bottom 20% of the country in terms of COVID-19 vaccine coverage* according to socioeconomic and health variables. Brazil, 2021-2022.

|  | Children | Adolescent | Adult | Elderly |
| --- | --- | --- | --- | --- |
| **Income concentration (GINI index)** |  |  |  |  |
| Quartile 1 (lower income concentration) | 10.8 | 13.8 | 8.8 | 7.9 |
| Quartile 2 | 14.3 | 14.9 | 14.8 | 13.2 |
| Quartile 3 | 21.2 | 20.1 | 23.0 | 25.3 |
| Quartile 4 (higher income concentration) | 37.0 | 33.8 | 36.8 | 36.5 |
| **Expected years of schooling at age 18** |  |  |  |  |
| Quartile 4 (more years of schooling) | 8.8 | 11.4 | 5.3 | 9.0 |
| Quartile 2 | 17.3 | 18.8 | 16.2 | 16.7 |
| Quartile 3 | 21.5 | 20.6 | 21.7 | 19.9 |
| Quartile 1 (less years of schooling) | 32.1 | 29.1 | 36.6 | 34.2 |
| **Proportion of black population** |  |  |  |  |
| Quartile 1 (less black population) | 9.3 | 17.1 | 3.6 | 5.4 |
| Quartile 2 | 9.3 | 9.8 | 7.1 | 8.5 |
| Quartile 3 | 24.2 | 20.1 | 23.0 | 25.0 |
| Quartile 4 (more black population) | 37.2 | 33.1 | 46.4 | 41.2 |
| **Per capita public spending on health** |  |  |  |  |
| Quartile 4 (higher expenditure) | 11.9 | 13.3 | 7.6 | 11.0 |
| Quartile 2 | 16.4 | 16.0 | 16.5 | 17.2 |
| Quartile 3 | 20.4 | 19.9 | 23.1 | 23.0 |
| Quartile 1 (lower expenditure) | 31.3 | 30.8 | 32.7 | 28.9 |
| **Nurses and doctors per 1,000 population** |  |  |  |  |
| Quartile 4 (more health professionals) | 13.5 | 16.4 | 7.5 | 7.8 |
| Quartile 2 | 18.2 | 17.1 | 15.1 | 15.6 |
| Quartile 3 | 19.5 | 19.3 | 22.0 | 22.5 |
| Quartile 1 (less health professionals) | 28.8 | 27.2 | 35.4 | 34.0 |
| **PHC ambulatory office per 1,000 population** |  |  |  |  |
| Quartile 4 (more offices) | 10.6 | 14.7 | 9.6 | 12.5 |
| Quartile 2 | 19.7 | 17.2 | 18.0 | 18.1 |
| Quartile 3 | 24.4 | 23.1 | 24.9 | 22.6 |
| Quartile 1 (less offices) | 25.3 | 25.0 | 27.6 | 26.8 |

\*: vaccination coverage analyzed in the epidemiological week in which the country reached 20% coverage in the respective age group.

The adjusted logistic regression analysis revealed that municipalities with lower education levels had higher chances of having the lower vaccination coverage (Table 3). Odds ratios ranged from 1.78 (95% CI 1.44-2.20) for children to 2.14 (95% CI 1.87-2.45) for adults in the quartile with the lowest education level, compared to the group with the highest expected years of study (Table 3). Additionally, municipalities with higher income concentration and a larger proportion of Black residents were more likely to have lower vaccination coverage. The odds ratio was 2.64 (95% CI 2.05-3.39) for the quartile with the highest Gini index in the elderly vaccination coverage analysis, and 9.67 (95% CI 6.89-13.58) for the quartile with the highest proportion of Black residents in the adult vaccination analysis.

**Table 3.** Adjusted model (logistic regression) of the association between lower COVID-19 vaccination coverage* and socioeconomic and health variables. Brazil, 2021-2022.

|  | Children | Adolescent | Adult | Elderly |
| --- | --- | --- | --- | --- |
| **Income concentration (GINI index)** | OR (CI<sub>95%</sub>) | OR (CI<sub>95%</sub>) | OR (CI<sub>95%</sub>) | OR (CI<sub>95%</sub>) |
| Quartile 1 (lower income concentration) | 1.00 | 1.00 | 1.00 | 1.00 |
| Quartile 2 | 1.09 (0.80-1.28) | 0.93 (0.74-1.16) | 0.99 (0.76-1.29) | 1.21 (0.93-1.57) |
| Quartile 3 | 1.23 (0.99-1.54) | 1.12 (0.90-1.38) | 1.12 (0.88-1.43) | 2.02 (1.59-2.57) |
| Quartile 4 (higher income concentration) | 2.07 (1.65-2.61) | 1.84 (1.48-2.30) | 1.52 (1.18-1.96) | 2.64 (2.05-3.39) |
| **Expected years of schooling at age 18** |  |  |  |  |
| Quartile 4 (more years of schooling) | 1.00 | 1.00 | 1.00 | 1.00 |
| Quartile 2 | 1.39 (1.08-1.78) | 1.58 (1.25-1.98) | 1.67 (1.24-2.26) | 0.99 (0.76-1.28) |
| Quartile 3 | 1.38 (1.06-1.78) | 1.58 (1.25-2.01) | 1.52 (1.12-2.06) | 0.79 (0.61-1.04) |
| Quartile 1 (less years of schooling) | 1.78 (1.37-2.30) | 1.97 (1.54-2.52) | 2.14 (1.59-2.90) | 1.14 (0.87-1.48) |
| **Proportion of black population** |  |  |  |  |
| Quartile 1 (less black population) | 1.00 | 1.00 | 1.00 | 1.00 |
| Quartile 2 | 0.86 (0.66-1.13) | 0.44 (0.35-0.56) | 1.71 (1.20-2.45) | 1.47 (1.08-2.01) |
| Quartile 3 | 1.95 (1.51-2.51) | 0.75 (0.59-0.94) | 4.47 (3.19-6.27) | 3.83 (2.84-5.15) |
| Quartile 4 (more black population) | 2.76 (2.12-3.58) | 1.13 (0.90-1.44) | 9.67 (6.89-13.58) | 5.91 (4.37-7.99) |
| **Per capita public spending on health** |  |  |  |  |
| Quartile 4 (higher expenditure) | 1.00 | 1.00 | 1.00 | 1.00 |
| Quartile 2 | 1.07 (0.85-1.34) | 1.05 (0.84-1.31) | 1.49 (1.15-1.95) | 1.06 (0.84-1.35) |
| Quartile 3 | 1.12 (0.89-1.41) | 1.21 (0.97-1.50) | 1.66 (1.28-2.16) | 1.14 (0.90-1.44) |
| Quartile 1 (lower expenditure) | 1.57 (1.25-1.97) | 1.72 (1.39-2.13) | 1.92 (1.48-2.50) | 1.10 (0.87-1.41) |
| **Nurses and doctors per 1,000 population** |  |  |  |  |
| Quartile 4 (more health professionals) | 1.00 | 1.00 | 1.00 | 1.00 |
| Quartile 2 | 1.04 (0.84-1.30) | 0.85 (0.69-1.05) | 1.40 (1.07-1.84) | 1.73 (1.34-2.24) |
| Quartile 3 | 0.85 (0.67-1.06) | 0.82 (0.66-1.02) | 1.61 (1.24-2.10) | 2.12 (1.64-2.73) |
| Quartile 1 (less health professionals) | 1.10 (0.88-1.38) | 1.00 (0.81-1.24) | 2.40 (1.85-3.12) | 3.12 (2.42-4.02) |
| **PHC ambulatory office per 1,000 population** |  |  |  |  |
| Quartile 4 (more offices) | 1.00 | 1.00 | 1.00 | 1.00 |
| Quartile 2 | 1.62 (1.29-2.04) | 1.02 (0.82-1.26) | 1.33 (1.04-1.71) | 1.03 (0.82-1.30) |
| Quartile 3 | 1.64 (1.30-2.06) | 1.24 (1.00-1.53) | 1.34 (1.05-1.71) | 0.95 (0.75-1.19) |
| Quartile 1 (less offices) | 1.63 (1.29-2.06) | 1.32 (1.07-1.64) | 1.46 (1.14-1.87) | 1.14 (0.90-1.43) |

\*: low vaccination coverage: to be among the 20% of municipalities with the lowest vaccination coverage in the epidemiological week in which the country reached a coverage equal to 20% in the respective age group.

Apart from the elderly, municipalities with lower per capita health expenditure had higher chances of having the lowest vaccination coverage. Odds ratios were 1.57 (CI95% 1.25-1.97) for children and 1.92 (CI95% 1.48-2.50) for adults. Furthermore, a lower number of primary health care ambulatory offices per 1,000 inhabitants increased the likelihood of municipalities being in the lowest vaccination coverage group, with odds ratios of 1.63 (95% CI 1.29-2.06) for children and 1.32 (95% CI 1.07-1.64) for adolescents. Lastly, a lower ratio of physicians and nurses to the population increased the chances of municipalities having poor vaccination coverage for adults (+140%) and the elderly (+212%). Unadjusted model values can be found in Supplementary Table 2.

The proportion of municipalities with the lowest vaccination coverage in Brazil varied significantly across different states and regions of the country (Supplementary Table 3). In the North region, 73.5% of municipalities were among the 1,114 municipalities with the lowest proportion of vaccinated population, while the percentage was much lower in the Southeast (6.1%) and the South regions (10.6%). This pattern was observed across all age groups. When comparing states, almost all municipalities in Roraima, a rural state with a considerable indigenous population, had the lowest vaccination coverage for children, adolescents, and adults. In contrast, only 0.8% of cities in the richest state, São Paulo, were in the lowest vaccine coverage. Figure 2 shows the spatial distribution of municipalities that make up the group of 20% with the lowest vaccination coverage in Brazil. Deep regional inequality is expressed with a higher proportion of municipalities with low coverage in the North region, the northern part of the Midwest, and the eastern part of the Southeast region.

**Figure 2.**
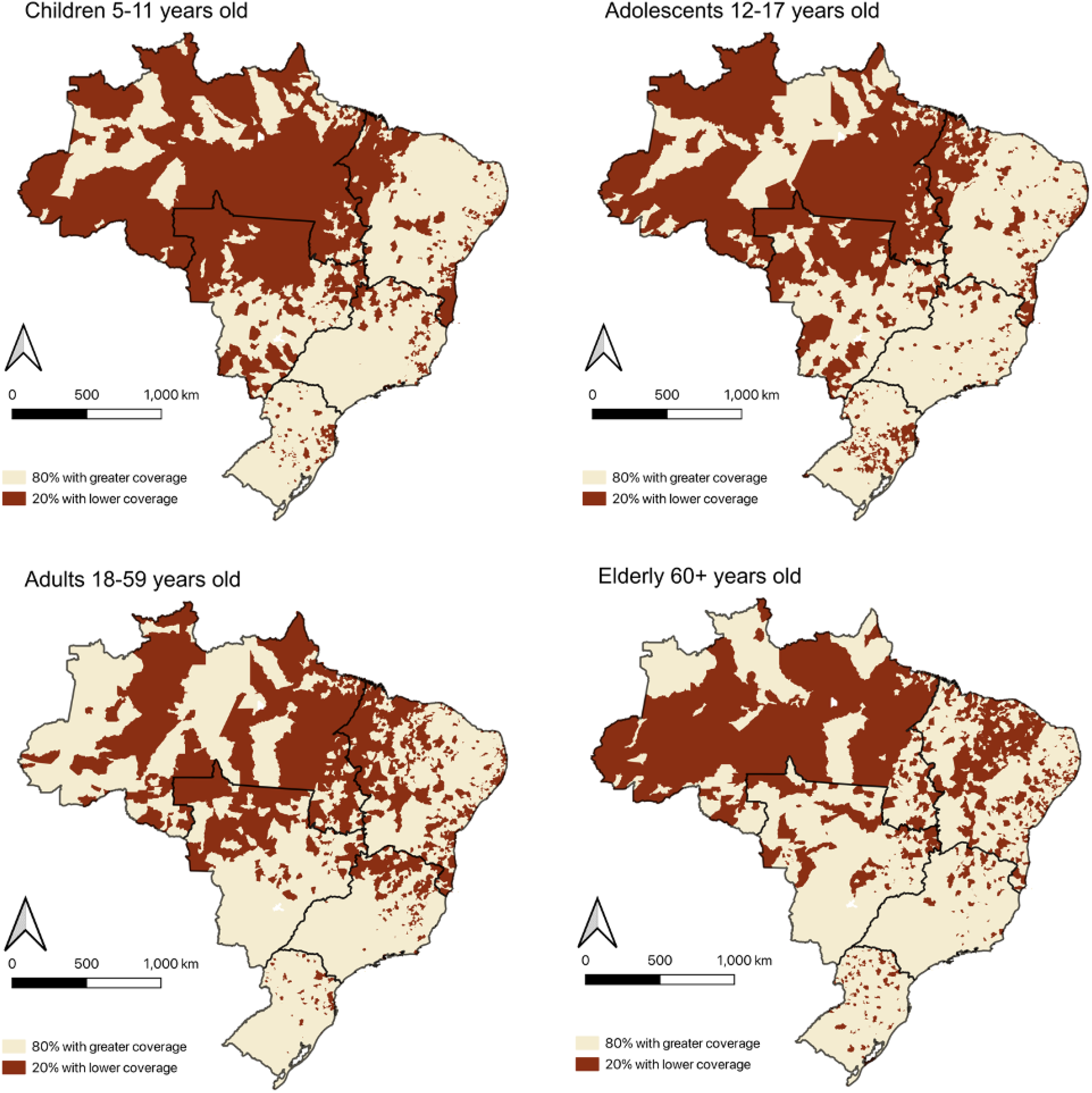
Spatial distribution of municipalities that make up the groups of 20% with the lowest and 80% with the highest covid-19 vaccination coverage*. Brazil, 2019-2020. *: vaccination coverage analyzed in the epidemiological week in which the country reached 20% coverage in the respective age group.

## Discussion

Our study has identified several significant findings related to COVID-19 vaccination coverage in Brazilian municipalities. Firstly, municipalities with lower vaccination coverage at the start of the campaign exhibited a smaller increase in coverage over the following months, resulting in increasing inequalities in all age groups as the campaign progressed. Secondly, municipalities with a higher proportion of Black population and poorer socioeconomic and healthcare indicators were more likely to have the lowest vaccination coverage. Finally, there are significant regional inequalities, with the worst vaccination coverage found in the North region, as well as in areas of the Midwest and Northeast.

The COVID-19 vaccination campaign has revealed profound inequalities between countries in access to vaccines since its inception. As countries began their vaccination campaigns, inequalities started to be observed within these countries as well.^22-25^ The results from Brazil align with these previous findings. Several possible explanatory hypotheses can be cited to explain such results. Firstly, inadequate monitoring of regional and socioeconomic inequalities in vaccination by the federal government and states may have contributed to the persistence of disparities in immunization coverage. In fact, the Brazilian government’s actions during the pandemic did not prioritize tackling COVID-19-related inequalities, despite the country’s status as one of the most unequal nations on the planet. Previous analyses have revealed that the federal government’s actions in 2021/2022 were characterized by delayed procurement, denialism, conspiracy theories, and vaccine skepticism, creating a scenario of intentional national disarticulation in the vaccination campaign.^26^

Our study adds a novel perspective by highlighting that the group of municipalities with the worst vaccination coverage at the beginning of the campaign remained behind, with fewer people immunized at the end of the analyzed period. This means that not only does regional inequality exist in vaccination coverage, but it has also persisted or even increased as the country progressed in its vaccination campaign. Although some state governments attempted to carry out joint actions^27^, their pandemic response strategies did not prioritize monitoring and tackling inequalities.^28^ Without sufficient concern for possible disparities and political will to address them, no information was produced to help guide equitable actions. As a result, there seems to have been a lack of technical and financial support for municipalities with worse indicators, which may have contributed to the persistence of inequalities in immunization coverage.

The municipalities with the lowest vaccination coverage that were left behind share geographic, demographic, socioeconomic, and health service similarities. Municipalities with a history of lower investment in health, less access to health services, and fewer healthcare professionals may face greater difficulties in carrying out a successful covid-19 vaccination campaign. For example, these municipalities may have a shortage of vaccination posts, making it more challenging for the population to access vaccines. In addition, the organizational capacity to manage a complex vaccination campaign may be limited in these municipalities. Physical infrastructure and work processes can also pose a challenge for these municipalities. For instance, the lack of adequate refrigeration facilities could limit the ability to store and transport vaccines safely. Furthermore, healthcare professionals in these municipalities may have fewer resources and training opportunities, which could affect their ability to administer vaccines safely and effectively, and register and keep immunization records up to date.^29-30^ Studies have found that people with higher income and education tend to show less hesitancy towards getting vaccinated against COVID-19.^31-32^ In Brazil, Moore et al.^33^ reported that individuals with less than nine years of education and a monthly income of less than US$789 were 31% and 13% more likely to express vaccine hesitancy, respectively. In Brazil vaccine-related information was often politicized, and social media platforms were rife with false and conspiratorial news, leading to negative impacts on vaccination decisions.^17^ Furthermore, populations with limited access to healthcare professionals and reliable sources of information were more susceptible to misinformation, further exacerbating the inequalities in vaccination coverage. Therefore, it is imperative that governments and social media companies take necessary actions to prevent the spread of misinformation and protect the lives of people. However, it is likely that the material and structural conditions of cities and the health services available to people had the greatest impact on vaccination in Brazil. Compared to other countries, vaccine hesitancy was low in Brazil and did not vary substantially between regions, being slightly lower in the North.^33-34^ And it was precisely in this region where the highest concentration of municipalities with low vaccination coverage was observed. A study by Miclos et al.^35^ evaluated the efficacy, relevance, and effectiveness of Primary Health Care (PHC) actions in Brazilian municipalities and found that the proportion of municipalities with “satisfactory” indicators was four times lower in the North compared to the South of the country. Additionally, Northern Region residents are less satisfied with the PHC services.^36^ These findings suggest that the availability and quality of health services are critical factors in determining the success of vaccination efforts.

The present study has limitations. The resident population values used for vaccine coverage calculations are estimates calculated from projections that considered the last census with data available in Brazil (2010). Likewise, socioeconomic data come from the same 2010 census, which is the last available year. The risk of inaccuracy resulting from the gap between the present day and the last census, however, was minimized by grouping municipalities according to sociodemographic and health indicators in many of the analyses in the present study. Furthermore, both population and socioeconomic data are official data from the Brazilian government and were made available by the Ministry of Health. During the covid-19 vaccination campaign, the SI-PNI faced limitations in recording data on the administered doses. There were delays in recording doses and problems with data integrity. To minimize the impact of any notification delays on coverage calculations, we used the database updated in March 2023, which covers all doses administered until December 2022.

## Data Availability

All data produced are available online at https://github.com/covid19br/dados-vacinas/tree/main/municipios and https://datasus.saude.gov.br/

## Ethics

All analyzed data is public and anonymized, and there was no need for ethical research committee approval.

## Acknowledgments

To Brazilian National Council for Scientific and Technological Development CNPq) for the grant for research productivity to Antonio Fernando Boing (312987/2021-8).

