## Supplemental material for "The Brazilian vaccine divide: how some municipalities are being left behind in the Covid-19 vaccine coverage"

Supplementary Table 1 – General description of the exploratory variables and covid-19 vaccine coverage. Brazil, 2021-2022.

| **Country characteristics** |  |
| --- | --- |
| Expected years of schooling at age 18* | 9.5 years |
| Proportion of Black residentes* | 50.9% |
| Gini index* | 0.600 |
| Gasto per capita em saúde** | R$908.4 |
| Enfermeiros e médicos por 1.000 habitantes*** | 3.50 |
| Consultórios em ambulatórios por 1.000 habitantes*** | 0.78 |
| **Percentage with first doseꟸ** |  |
| 5-11 years old | 69.5% |
| 12-17 years old | 95.1% |
| 18-59 years old | 97.4% |
| 60+ years old | 98.4% |
| **Percentage with second doseꟸ** |  |
| 5-11 years old | 46.2% |
| 12-17 years old | 75.8% |
| 18-59 years old | 86.3% |
| 60+ years old | 94.5% |
| **Percentage with third dose (booster)ꟸ** |  |
| 12-17 years old | 24.8% |
| 18-59 years old | 52.3% |
| 60+ years old | 79.7% |

*: According to 2010 demographic census; **: Year 2020; ***: January 2021; **ꟸ:** December 2022

Supplementary Table 2 - Crude model (logistic regression) of the association between lower COVID-19 vaccine coverage* and socioeconomic and health variables. Brazil, 2021-2022.

|  | Children | Adolescent | Adult | Elderly |
| --- | --- | --- | --- | --- |
| **Income concentration (GINI index)** | OR (CI_95%_) | OR (CI_95%_) | OR (CI_95%_) | OR (CI_95%_) |
| Quartile 1 (lower income concentration) | 1.00 | 1.00 | 1.00 | 1.00 |
| Quartile 2 | 1.39 (1.11-1.74) | 1.09 (0.88-1.34) | 1.81 (1.43-2.28) | 1.77 (1.39-2.27) |
| Quartile 3 | 2.24 (1.83-2.74) | 1.56 (1.29-1.89) | 3.12 (2.53-3.86) | 3.94 (3.17-4.90) |
| Quartile 4 (higher income concentration) | 4.87 (3.99-5.95) | 3.18 (2.63-3.84) | 6.07 (4.91-7.50) | 6.70 (6.70-8.34) |
| **Expected years of schooling at age 18** |  |  |  |  |
| Quartile 4 (more years of schooling) | 1.00 | 1.00 | 1.00 | 1.00 |
| Quartile 2 | 2.16 (1.71-2.72) | 1.80 (1.45-2.23) | 3.45 (2.62-4.54) | 2.02 (1.60-2.55) |
| Quartile 3 | 2.82 (2.25-3.54) | 2.02 (2.23-2.49) | 4.96 (3.80-6.49) | 2.52 (2.01-3.16) |
| Quartile 1 (less years of schooling) | 4.87 (3.92-6.05) | 3.20 (2.61-3.91) | 10.32 (7.97-13.38) | 5.25 (4.24-6.51) |
| **Proportion of black population** |  |  |  |  |
| Quartile 1 (less black population) | 1.00 | 1.00 | 1.00 | 1.00 |
| Quartile 2 | 1.01 (0.78-1.30) | 0.52 (0.42-0.66) | 2.06 (1.45-2.91) | 1.63 (1.21-2.20) |
| Quartile 3 | 3.13 (2.51-3.89) | 1.22 (1.00-1.47) | 8.01 (5.88-10.91) | 5.83 (4.48-7.58) |
| Quartile 4 (more black population) | 5.79 (4.69-7.16) | 2.40 (2.00-2.86) | 23.22 (17.18-31.38) | 12.31 (9.53-15.90) |
| **Per capita public spending on health** |  |  |  |  |
| Quartile 4 (higher expenditure) | 1.00 | 1.00 | 1.00 | 1.00 |
| Quartile 2 | 1.45 (1.17-1.79) | 1.24 (1.01-1.54) | 2.40 (1.88-3.06) | 1.68 (1.35-2.09) |
| Quartile 3 | 1.89 (1.54-2.33) | 1.62 (1.32-1.98) | 3.65 (2.89-4.61) | 2.42 (1.96-2.98) |
| Quartile 1 (lower expenditure) | 3.37 (2.76-4.10) | 2.91 (2.40-3.52) | 5.91 (4.71-7.42) | 3.28 (2.68-4.03) |
| **Nurses and doctors per 1,000 population** |  |  |  |  |
| Quartile 4 (more health professionals) | 1.00 | 1.00 | 1.00 | 1.00 |
| Quartile 2 | 1.42 (1.16-1.74) | 1.05 (0.86-1.28) | 2.20 (1.72-2.82) | 2.18 (1.71-2.78) |
| Quartile 3 | 1.56 (2.27-1.90) | 1.22 (1.01-1.48) | 3.50 (2.76-4.44) | 3.41 (2.71-4.31) |
| Quartile 1 (less health professionals) | 2.59 (2.14-3.14) | 1.91 (1.59-2.30) | 6.78 (5.40-8.52) | 6.07 (4.84-7.60) |
| **PHC ambulatory office per 1,000 population** |  |  |  |  |
| Quartile 4 (more offices) | 1.00 | 1.00 | 1.00 | 1.00 |
| Quartile 2 | 2.06 (1.66-2.55) | 1.20 (0.98-1.47) | 2.07 (1.65-2.59) | 1.55 (1.25-1.91) |
| Quartile 3 | 2.72 (2.20-3.35) | 1.74 (1.43-2.11) | 3.14 (2.53-3.90) | 2.05 (1.67-2.51) |
| Quartile 1 (less offices) | 2.84 (2.31-3.50) | 1.94 (1.60-2.34) | 3.60 (2.91-4.46) | 2.56 (2.10-3.12) |

*: low vaccination coverage: to be among the 20% of municipalities with the lowest vaccination coverage in the epidemiological week in which the country reached a coverage equal to 20% in the respective age group; OR: odds ratio; CI_95%_: confidence interval.

Supplementary Table 3 - Absolute number and proportion of municipalities from each Brazilian state that are among the 20% of the country with the lowest vaccination coverage in each age group*. Brazil, 2021-2022.

|  | Children | Adolescent | Adult | Elderly |
| --- | --- | --- | --- | --- |
| **North** | **330 (73.5)** | 309 (68.8) | 265 (59.0) | 240 (53.4) |
| Rondônia | 50 (96.2) | 36 (69.2) | 20 (38.5) | 16 (30.8) |
| Acre | 20 (90.9) | 14 (63.6) | 6 (27.3) | 19 (86.4) |
| Amazonas | 35 (56.4) | 40 (48.4) | 18 (29.0) | 44 (71.0) |
| Roraima | 14 (93.3) | 15 (100.0) | 13 (86.7) | 3 (20.0) |
| Pará | 90 (62.9) | 102 (71.3) | 107 (74.8) | 118 (82.5) |
| Amapá | 11 (68.8) | 8 (50.0) | 14 (87.5) | 1 (6.25) |
| Tocantins | 110 (79.1) | 104 (74.8) | 87 (62.6) | 39 (28.1) |
| **Northeast** | **372 (20.7)** | **359 (20.0)** | 517 (28.8) | 610 (34.0) |
| Maranhão | 170 (78.4) | 142 (65.4) | 151 (69.6) | 59 (27.2) |
| Piauí | 4 (1.8) | 8 (3.6) | 96 (42.9) | 183 (81.7) |
| Ceará | 6 (3.3) | 9 (4.9) | 29 (15.8) | 134 (72.8) |
| Rio Grande do Norte | 11 (6.6) | 17 (10.2) | 11 (6.6) | 19 (11.4) |
| Paraíba | 11 (4.9) | 17 (7.6) | 29 (13.0) | 15 (6.7) |
| Pernambuco | 25 (13.5) | 29 (15.7) | 38 (20.5) | 49 (26.5) |
| Alagoas | 41 (40.2) | 53 (52.0) | 31 (30.4) | 34 (33.3) |
| Sergipe | 4 (5.3) | 4 (5.3) | 13 (17.3) | 12 (16.0) |
| Bahia | 100 (24.0) | 80 (19.2) | 119 (28.5) | 105 (25.2) |
| **Southeast** | **101 (6.1)** | **79 (4.7)** | 167 (10.0) | 53 (3.2) |
| Minas Gerais | 79 (9.3) | 53 (6.2) | 158 (18.5) | 48 (5.6) |
| Espírito Santo | 4 (5.1) | 7 (9.0) | 1 (1.3) | - |
| Rio de Janeiro | 18 (19.6) | 14 (15.2) | 8 (8.7) | 5 (5.4) |
| São Paulo | - | 5 (0.8) | - | - |
| **South** | **126 (10.6)** | **240 (20.2)** | 64 (5.4) | 115 (9.7) |
| Paraná | 14 (3.5) | 27 (6.8) | 25 (6.3) | 64 (16.0) |
| Santa Catarina | 69 (23.6) | 101 (34.5) | 33 (11.3) | 17 (5.8) |
| Rio Grande do Sul | 43 (8.7) | 112 (34.5) | 6 (1.2) | 34 (6.8) |
| **Midwest** | **184 (39.5)** | **126 (27.0)** | 101 (21.7) | 95 (20.4) |
| Mato Grosso do Sul | 25 (32.0) | 21 (26.9) | - | - |
| Mato Grosso | 95 (67.4) | 71 (50.4) | 65 (46.1) | 37 (26.2) |
| Goiás | 64 (26.0) | 34 (13.8) | 36 (14.6) | 58 (23.6) |

*: vaccination coverage analyzed in the epidemiological week in which the country reached 20% coverage in the respective age group.
